# Social Economic Inequality of Oral Health among U.S. Residents from 1999 to 2023 and International Comparison: A Multilevel Analysis Based on NHANES and WHO Oral Health Data

**DOI:** 10.64898/2025.12.10.25339882

**Authors:** Zhou Chengke

## Abstract

**Objective:** To investigate socioeconomic inequalities in oral health across global and national contexts, to quantify income related gradients in oral health among United States adults, and to elucidate the mechanisms through which socioeconomic status (SES) shapes oral health, with a focus on access to dental care and the potential role of state Medicaid policies.

**Design:** Multi level observational study combining macro level global and national comparisons with micro level individual data and meso level state policy analysis.

**Setting:** United States nationally representative surveys and publicly available international data.

**Participants:** Adults aged 20 years and older in the National Health and Nutrition Examination Survey (NHANES) 1999–2023 (pooled cross sectional samples), and state level adult populations from the Behavioral Risk Factor Surveillance System (BRFSS) 2011–20XX.

**Main outcome measures:** Individual outcomes were the Decayed Missing Filled Teeth index (DMFT) and self rated oral health (good versus fair or poor). State level outcomes were the prevalence of past year dental visits, any permanent tooth loss, and complete edentulism among adults aged 65 years and older. SES was measured by the poverty income ratio (PIR) and education. Mediators included annual dental visits, unmet dental need, and health behaviours. State policy variables included indicators of Medicaid expansion and adult Medicaid dental benefit generosity.

**Results:** In NHANES, higher PIR and higher education were independently associated with lower DMFT and higher odds of self rated good oral health across all age groups. PIR coefficients for DMFT were approximately -0.25 (20–44 years), -0.79 (45–64 years), and -1.07 (65 years and older), while the corresponding odds ratios for self rated good oral health were about 1.48, 1.46, and 1.33. Predicted probabilities of self rated good oral health increased monotonically from PIR quartile 1 to quartile 4 in every age group. Income related concentration indices indicated that caries and tooth loss were concentrated among low income adults (CI for DMFT about -0.105), whereas good oral health was concentrated among higher income adults (CI about 0.094). The Slope Index of Inequality suggested that moving from the lowest to highest income rank was associated with an average reduction of about 2.48 affected teeth, and the Relative Index of Inequality indicated approximately eight fold higher odds of reporting good oral health in the highest versus lowest income ranks. Oaxaca Blinder decomposition showed that adults in the highest income quartile had on average 1.31 fewer affected teeth than those in the lowest quartile; about one quarter of this gap was explained by observed variables, with access to dental care (annual visits and unmet need) accounting for roughly two thirds of the explained component. In state level difference in differences models with state and year fixed effects and state clustered standard errors, Medicaid expansion and adult dental benefit generosity were not associated with large, statistically significant changes in aggregate dental visit rates, any tooth loss, or edentulism among older adults.

**Conclusions:** Marked socioeconomic inequalities in oral health persist among United States adults, particularly in midlife, and are strongly linked to differences in access to dental care and socially patterned behaviours. State level Medicaid expansion and adult dental benefit policies, as measured here, did not produce large detectable changes in aggregate tooth loss outcomes, consistent with the highly cumulative nature of these endpoints and the dilution of effects in whole state populations. Expanding dental coverage remains necessary to reduce financial barriers and unmet need, but is unlikely to eliminate oral health inequalities without broader policies that address underlying income and education inequalities and other social determinants of health. These findings have implications for the integration of oral health into universal health coverage agendas in the United States, China, and other countries.

## 1 Introduction

### 1.1 Research Background: Global and National Inequality

Oral health is a vital component of overall health and well being. The World Health Organization defines oral health as a state free from chronic oral and facial pain, oral and pharyngeal cancers, oral infection and sores, periodontal disease, tooth decay, tooth loss, and other disorders that limit an individual’s capacity in biting, chewing, smiling, speaking, and psychosocial functioning. Untreated dental caries, severe periodontal disease, and edentulism are among the most prevalent non communicable conditions worldwide and impose substantial financial and social burdens on individuals, families, and health systems.

Despite this burden, the global distribution of oral health resources and outcomes remains highly unequal. High income countries generally have more dentists, greater use of preventive care, and lower levels of untreated disease, whereas many low and middle income countries face high disease burdens, scarce oral health workforce, and large gaps in access to basic dental treatment. Within countries, oral health outcomes often follow a clear socioeconomic gradient: people with lower income, less education, and more precarious employment tend to have higher rates of caries, tooth loss, and impaired oral function. This pattern has been described in many settings, including the United States, European countries, and China.

In China, successive national oral health surveys have documented persistent and in some cases widening disparities in caries, periodontal disease, and tooth loss across urban and rural areas, regions, and education and income groups. Although national strategies such as “Healthy China 2030” and school based oral health promotion have raised awareness and improved preventive care among some population groups, coverage of restorative and prosthodontic services through basic health insurance remains limited and uneven, and many adults still face substantial out of pocket payments for dental care. Similar challenges exist in other middle and high income countries where oral health services are only partially included in universal health coverage arrangements.

In the United States, a large body of research using NHANES and other datasets has shown pronounced socioeconomic inequalities in oral health. Low income and low education adults have higher DMFT scores, fewer functional teeth, worse self rated oral health, and higher levels of pain and functional limitation. Financial barriers to care, lack of dental insurance, irregular dental visiting patterns, and socially patterned behaviours such as smoking, unhealthy diet, and short sleep all contribute to these inequalities. At the same time, the United States relies heavily on private dental insurance and out of pocket payments, with public coverage for adults largely limited to Medicaid and highly variable across states. This creates a fragmented landscape in which both social and policy determinants shape oral health outcomes ^[3,4]^.

### 1.2 Theoretical framework: The mechanisms of health inequity

Understanding how socioeconomic status (SES) translates into oral health inequalities requires an explicit theoretical framework. The social determinants of health literature proposes several interrelated pathways. First, **material pathways** reflect the direct impact of income and wealth on the ability to purchase nutritious food, maintain good living conditions, and afford transportation and dental care. Higher SES reduces exposure to material hardship and lowers financial barriers to early detection and treatment of oral disease.

Second, **behavioural and cultural pathways** capture the fact that educational attainment and social position influence health literacy, beliefs, and self efficacy. Individuals with higher SES are more likely to adopt and maintain favourable behaviours such as twice daily toothbrushing with fluoride toothpaste, use of dental floss, non smoking, moderate sugar intake, and regular preventive dental visits. These behaviours can reduce the incidence and progression of oral disease.

Third, **psychosocial pathways** emphasise the role of chronic stress, job insecurity, discrimination, and lack of control in daily life. Chronic psychosocial stress can impair immune function, increase inflammatory responses, and contribute indirectly to oral disease through coping behaviours such as smoking, alcohol use, and poor sleep. Fourth, **institutional and service pathways** reflect the way SES shapes an individual’s position in the health system. People with higher income and stable employment are more likely to have comprehensive dental insurance, to live in areas with more providers, and to be able to take time off work for dental appointments. In contrast, low income adults often face a combination of lack of coverage, provider shortages, long waiting times, and rigid work schedules.

These pathways do not operate in isolation. They intersect with geographical differences (for example urban rural divides and regional resource allocation), physical environments (such as water fluoridation or endemic fluorosis), and policy environments (such as whether dental care is included in basic health insurance or universal health coverage). In both the United States and China, oral health is shaped by a combination of **structural socioeconomic inequalities** and **health system design**, including the scope and generosity of publicly financed dental benefits.

### 1.3 Research Significance and Purpose

The association between SES and oral health has been well documented, but important gaps remain. Many previous studies have described correlations between income, education, and oral health outcomes, yet relatively few have combined: (1) rigorous measurement of income related inequality using indices such as the concentration index (CI), Slope Index of Inequality (SII), and Relative Index of Inequality (RII); (2) formal mediation and decomposition analyses to identify the contribution of access to dental care and health behaviours to SES gradients; and (3) explicit consideration of how state level policy environments, such as Medicaid expansion and adult dental benefit generosity, may modify or fail to modify these inequalities. In addition, evidence from the United States is seldom discussed in parallel with findings from China and other countries that are seeking to integrate oral health into universal health coverage.

Against this background, the present study has four main objectives. First, using NHANES 1999–2023, we describe and quantify SES gradients in both objective (DMFT) and subjective (self rated oral health) outcomes among United States adults, including age stratified analyses across the adult life course. Second, we compute CI, SII, and RII to summarise income related inequalities in oral health and to compare their magnitude across age groups. Third, we examine potential mechanisms by conducting mediation analysis and Oaxaca Blinder decomposition to assess the roles of access to dental care, unmet dental need, and health behaviours in explaining income related gaps in DMFT and self rated oral health. Fourth, we construct a state year panel of Medicaid expansion status and adult dental benefit generosity, link it to BRFSS state level oral health indicators, and use difference in differences models to explore whether recent changes in Medicaid policies are associated with shifts in aggregate dental visit rates and tooth loss. By integrating micro level SES gradients, meso level policy analysis, and global and Chinese contextual evidence, we aim to provide a more comprehensive picture of oral health inequalities and their policy implications.

## 2. Data and Methods

### 2.1 Data Sources

NHANES (Microdata): This study utilized continuous-cycle public data from NHANES spanning 1999–2000 to 2021–2023 (data sourced from the Demographics/Dietary/Examination/Lab/Questionnaire module). NHANES is a nationally representative, stratified, multi-stage probability sampling survey conducted among demilitarized and non-institutionalized U.S. populations. The analysis included all data-complete adult participants (N=…) and performed subgroup analyses for adolescents (12–19 years old).

WHO (macro data): The macro background analysis used publicly available estimates from the WHO Global Oral Health Database, the Global Burden of Disease (GBD) study, and national indicators from the World Bank (e.g. GDP per capita, health spending as a percentage of GDP, etc.).

### 2.2 Variable and Index Construction

#### 2.2.1 Variable and Indicator Construction (NHANES)

##### 2.1.1.1. Outcomes

A dual-outcome model was used to capture both objective clinical measures and subjective health perceptions.

(1) DMFT (Decayed Missing Filled Total) is a continuous variable calculated from the tooth position field in the Oral Health Examination (OHXDEN) module, which counts and aggregates decayed, missing, and filled teeth. As the gold standard for assessing lifetime caries burden, DMFT quantifies the cumulative impact by measuring the total number of teeth that are untreated (Decayed), lost (Missing), or filled (Filled) due to caries.

(2) Self-assessment of oral health (self-good): Based on the questionnaire item "How would you rate your oral health status?" (oral_selfrate), responses of "excellent/very good/bad" are coded as 1 (self-assessed as good), while "average/poor" are coded as 0. This metric reflects a comprehensive evaluation of functional, aesthetic, and psychosocial satisfaction, serving as a key proxy for oral health-related quality of life (OHRQoL).

(3) Caries status (any_caries): A binary variable (yes/no) based on oral examination (OHX TC/SE/FL threshold judgment), used for behavioral factor analysis.

##### 2.1.1.2. Main exposure variables (SES)

(1) Education level (education_std) is categorized into: <High school (baseline reference), High school/GED, Some college/AA, College+. This indicator represents knowledge and cultural capital.

(2) Income (PIR): Household Income Poverty Ratio, a continuous variable. A lower PIR indicates greater poverty and represents material resources. Income quartiles (income_quartile_label, Q1-Q4) are used for descriptive analysis.

##### 2.1.1.3. Mediators & Behaviors

(1) Last dental visit in the past year (visit_last_year): coded as 1 (yes) for "within 6 months" or "within 1 year" based on the OHQ030 questionnaire (last dental visit), and 0 (no) otherwise, reflecting actual utilization of dental services.

(2) Unmet dental needs (unmet_need): coded as 1 for responses of ’yes’ to the OHQ770 questionnaire (last year needed but did not receive dental treatment), reflecting perceived barriers to healthcare accessibility.

(3) Reasons for unmet need: The descriptive analysis utilized variables from the OHQ780A-K scale, including ’cost’ (A), ’insurance’ (C), and ’time/busy’ (E, H, I), to identify the specific nature of the barriers.

(4) Short sleep (short_sleep): Based on the sleep questionnaire (sleep_hours), a sleep duration of less than 6 or 7 hours per night is defined as 1 (yes). It serves as a composite proxy for psychosocial stress and unhealthy lifestyle.

(5) Oral hygiene and beverages (Behaviors): including brushing frequency (brush_freq_num), frequency of consuming sugary drinks, tea consumption (tea_any), and caffeine intake (coffee_mg_day, caffeine_high_300mg).

##### 2.1.1.4. Covariates

(1) In the main effect and mediation models, covariates included age (continuous), sex, race/ethnicity, and behavioral variables. In the behavioral factors model, covariates included age, sex, race, PIR, and education.

(2) Note: The urban-rural variable (urban_rural_std) has been removed from all analyses as it is not available in the public data (URDHHLOC).

#### 2.2.2 Variable and Indicator Construction (WHO)

##### 2.2.2.1 Outcomes

untreated caries prevalence (1-9 years of age for deciduous teeth; 5+ years of age for permanent teeth), prevalence of severe periodontal disease (15+ years of age), prevalence of aphasia (20+ years of age), and sugar supply (g/person/day).

##### 2.2.2.2 Exposures

include geographical region (categorical dummy variable), per capita GDP (logarithmized), per capita dental expenditure (US), total dental expenditure (million US), dentist density (per 10,000 population), proportion of total health expenditure, tropical region status (binary), and coverage of related policies (binary).

### 2.3 Statistical Analysis Methods

All microanalyses were performed using Python software (statsmodels library) and took into account the complex sampling design of NHANES (weights, stratification, and clustering).

#### 2.3.1 NHANES Analysis Method

##### 2.3.1.1 Sampling design (NHANES)

WTMEC2YR was prioritized as the weighting factor (since the core outcome DMFT was derived from MEC examinations), with SDMVPSU for PSUs and SDMVSTRA for strata. Weight loss was backfilled to 1 (only when necessary).

##### 2.3.1.2 Descriptive analysis (NHANES)

Sample characteristics were described using weighted means (continuous variables) or proportions (categorical variables). Dental service accessibility indicators were stratified by SES (education, income quintiles) and trend charts were plotted.

##### 2.3.1.3 Single-factor analysis (NHANES)

ANOVA, t-test, and chi-square test were used to compare differences in DMFT or self-rated good/bad between education, income, race, and sleep duration groups.

##### 2.3.1.4 Behavioral Factor Analysis (NHANES)

To independently assess the association between health behaviors and caries risk (any_caries), a weighted logistic regression model was constructed using cluster-robust or HC1 standard errors. Principal component analysis (PCA) was performed on behavioral variables to extract PC1/PC2, which were used to explore the relationship between comprehensive behavioral patterns and caries risk.

##### 2.3.1.5 SES main effect model (NHANES)

A generalized linear model (GLM) was employed. For the continuous outcome DMFT, the weighted least squares (WLS) method was used. For the binary outcome (self-rated health), a weighted binary logistic regression was applied. All models utilized PSU-clustered robust standard errors (falling back to HC1 when PSU was insufficient) to ensure more reliable variance estimates.

##### 2.3.1.6 Mediation analysis (NHANES)

was conducted using the Product Method combined with Bootstrap (N=300) to estimate the indirect effect and its 95% confidence interval (CI). The pathways SES (PIR/Education) → M (Access/Sleep) → Y (DMFT/Self-good) were tested separately.

##### 2.3.1.7 Robustness and Nonlinearity Analysis (NHANES)

To ensure model reliability, we tested models with different covariate sets (basic, +behavior, +caffeine) and time windows (e.g., excluding 2005–2008 and 2021–2023). Restricted Cubic Spline (RCS) testing was used to examine nonlinear effects of PIR and age, capturing potential dose-response relationships.

#### 2.3.2 WHO analysis method

Regional differences in outcomes were analyzed using ANOVA or Kruskal-Wallis tests. Spearman ρcorrelation analysis was employed to examine the relationship between national economic indicators and outcomes. Multivariate linear regression or logistic regression was used to evaluate the associations between national economic, resource, and policy indicators with oral health outcomes, with regional stratified regression testing.

#### 2.3.3 Inequality indices (concentration index, SII and RII)

To formally quantify income-related inequalities in oral health, we calculated three standard summary indices across the full adult sample and within age strata (20–44, 45–64, and ≥65 years). First, we computed the concentration index (CI) for DMFT and self-rated oral health using the relative rank of the poverty–income ratio (PIR) in the national income distribution. DMFT is a “bad” outcome, for which a negative CI indicates that higher caries and tooth loss are concentrated among low-income groups; self-rated good oral health is a “good” outcome, for which a positive CI indicates that good health is concentrated among high-income groups.

Second, we estimated the Slope Index of Inequality (SII) and Relative Index of Inequality (RII) using regression models with PIR rank as the main exposure. For DMFT, SII was obtained from a linear regression of DMFT on PIR rank (ranging from 0 for the lowest income group to 1 for the highest), adjusted for age, sex and race/ethnicity. The SII coefficient represents the absolute difference in DMFT between the theoretical lowest and highest positions in the income hierarchy. For self-rated good oral health, we fitted logistic regression models of the log-odds of self-rated good on PIR rank to obtain an RII on the odds ratio scale, representing the relative difference (ratio) in the probability of good oral health between the highest and lowest income ranks. All indices were estimated using NHANES survey weights and robust standard errors that accounted for the complex sampling design.

#### 2.3.4 Oaxaca–Blinder decomposition of income-related gaps in DMFT

To further disentangle the mechanisms underlying SES inequalities, we applied a twofold Oaxaca–Blinder decomposition to the DMFT difference between adults in the lowest (Q1) and highest (Q4) PIR quartiles. The outcome was continuous DMFT, and the linear regression models included the following domains of explanatory variables:

(1) Access to dental care (annual dental visit, unmet dental need in the past year);
(2) Health behaviours (short sleep duration and selected behavioural indicators);
(3) Education (dummy variables for education level);
(4) Demographics (age in decades, sex); and
(5) Race/ethnicity (categorical indicator).

Separate regressions were estimated for Q1 and Q4 using survey-weighted linear models. The total mean difference in DMFT between Q1 and Q4 was decomposed into an “explained” component attributable to differences in the distribution of the explanatory variables across income groups (endowment effect) and an “unexplained” component reflecting differences in regression coefficients (coefficient effect), which may capture unmeasured factors and structural discrimination. We grouped individual variables into the above domains and summed their contributions to obtain domain-level contributions to the explained component. To avoid numerical artefacts (e.g. zero contributions due to matrix singularity), we ensured that the design matrix remained as a pandas DataFrame with float-typed columns and excluded redundant income variables (PIR and income quartiles were not simultaneously included).

#### 2.3.5 State-level Medicaid policy panel and difference-in-differences analysis

In a complementary state-level analysis, we constructed a state–year policy panel by merging BRFSS prevalence data on oral health outcomes with state Medicaid policy indicators. BRFSS state-level age-adjusted prevalence estimates were obtained from the CDC chronic disease indicators for 2011–2025. For each state and year, we extracted: (1) the proportion of adults who reported visiting a dentist or dental clinic for any reason within the past 12 months (dental_visit_rate); (2) the prevalence of any permanent tooth loss among adults (any_tooth_loss_rate); and (3) the prevalence of complete edentulism among adults aged ≥65 years (edentulism_65_rate).

Medicaid policy variables were derived from publicly available sources. The ACA Medicaid expansion indicator was coded using Kaiser Family Foundation (KFF) documentation on the timing of expansion decisions; for each state and year, MedicaidExpansion_st was set to 1 in years after the state implemented expansion and 0 otherwise. Adult Medicaid dental benefit generosity was coded using classifications from the Center for Health Care Strategies (CHCS), MACPAC compendia and CareQuest Institute reports. States were categorised as providing no or emergency-only coverage, limited coverage, or extensive coverage, and an ordinal variable DentalGenerosityLevel_st was constructed taking values 0, 1 and 2 respectively.

We then estimated difference-in-differences (DID) models of the form:

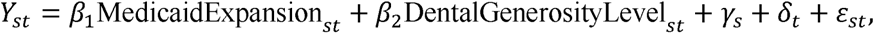

where *Y_St_* denotes each of the three state-level oral health outcomes in state sand year *t*; *γ_S_* and *δ_t_* are state and year fixed effects; and *ε_st_* is the error term. All models were estimated using ordinary least squares with standard errors clustered at the state level. The main analysis focused on dental_visit_rate as a proximal indicator of access to care, while any_tooth_loss_rate and edentulism_65_rate were examined as distal oral health outcomes. These state-level models are intended to complement, rather than replicate, the individual-level NHANES analyses, and to provide a rough assessment of whether recent changes in Medicaid coverage were associated with shifts in aggregate oral health indicators.

## 3. Results

### 3.1 NHANES Individual-level Results

#### 3.1.1 Description and trends: the huge gap in service accessibility

There is a strong SES gradient in both dental service utilization and unmet needs (see Figure X).

##### 3.1.1.1 Medical Utilization (OHQ030)

The overall population’s healthcare utilization rate over the past 12 months was approximately 63-65%. However, this average masks significant disparities: High-income (Q4) groups (e.g., 80% in 2013-2014) had significantly higher annual utilization rates than low-income (Q1) groups (approximately 52%). Similarly, highly educated (College+) groups (around 75% in 2019-2020) demonstrated substantially higher utilization rates compared to less educated groups (about 40%).

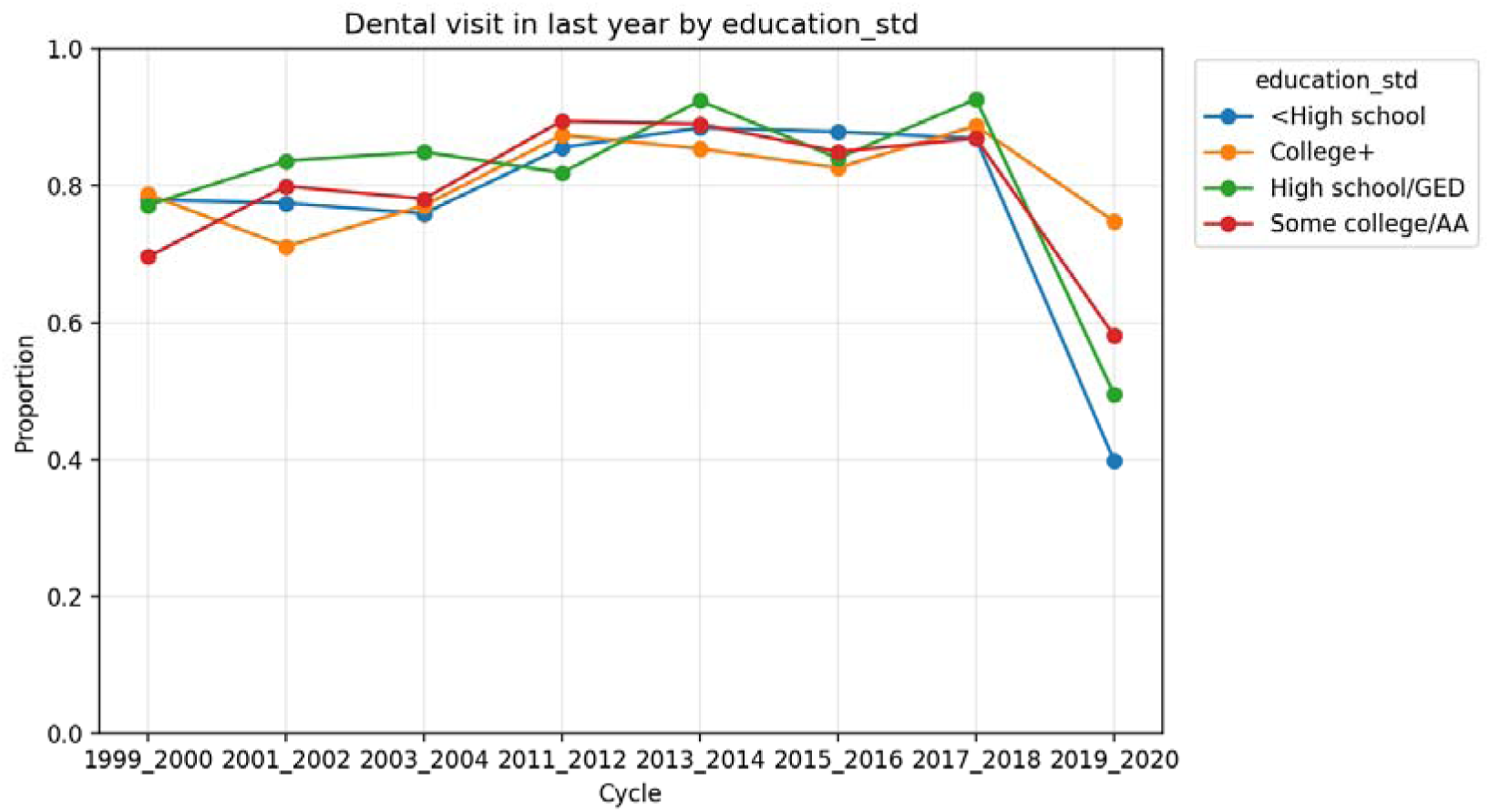

##### 3.1.1.2 Unmet need (OHQ770)

The proportion of the general population who do not have access to dental care is about 14-16%. Similarly, the proportion of low-income (Q1) people who report "need but do not have access to dental care" is very high (about 25-30%), almost five times higher than that of high-income (Q4) people (about 5-6%).

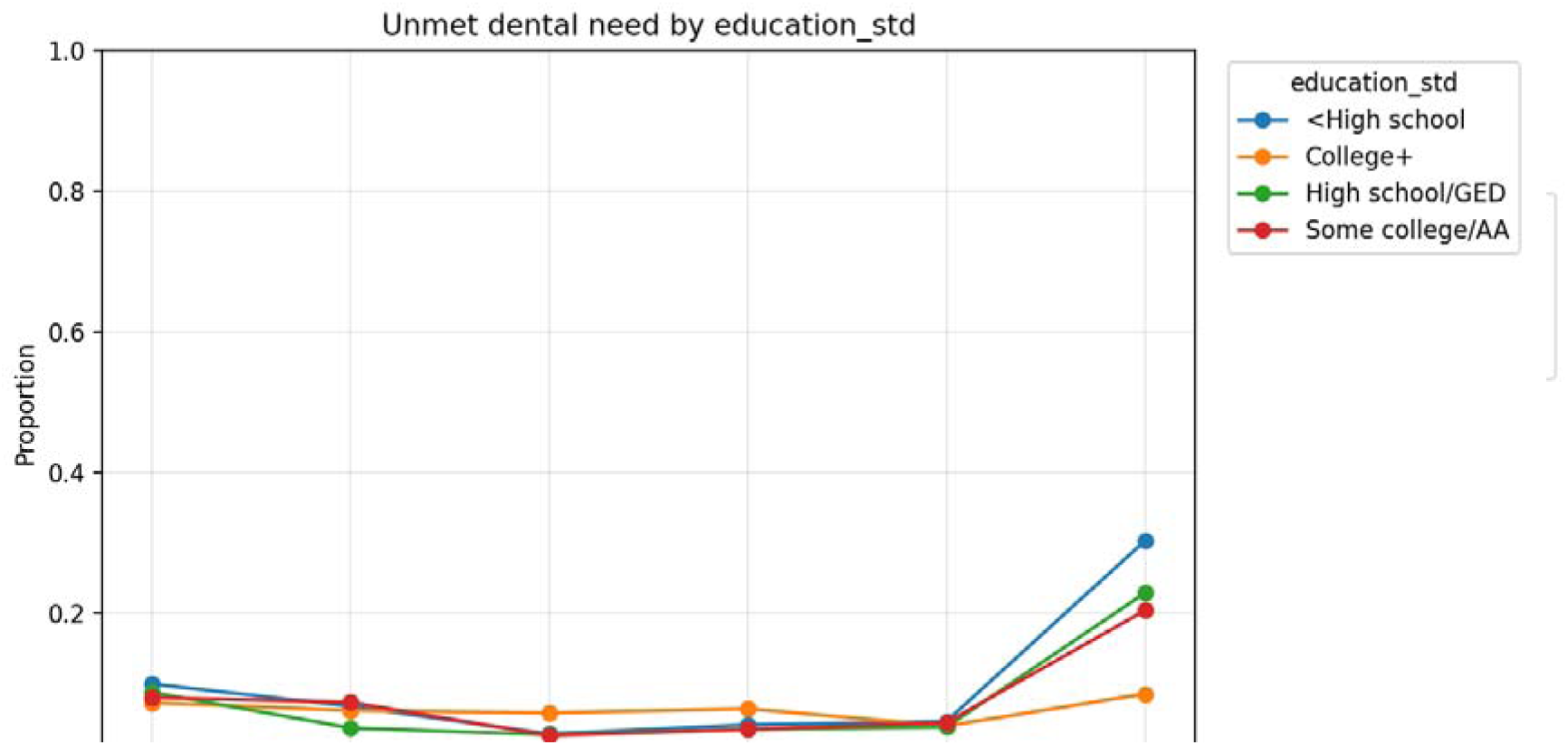

##### 3.1.1.3 Reasons for unmet needs (OHQ780A-K)

Among all respondents reporting unmet needs, "unaffordability (OHQ780A)" was the predominant cause, accounting for 70-80%. In low-income (Q1) groups, approximately 75-84% cited this as the reason, while the proportion was significantly lower in high-income (Q4) groups. "Insurance noncoverage (OHQ780C)" ranked second, with structural barriers like "time/busy/clinic inconvenience" being less common. These findings provide direct evidence that "economic barriers" serve as the core mechanism.

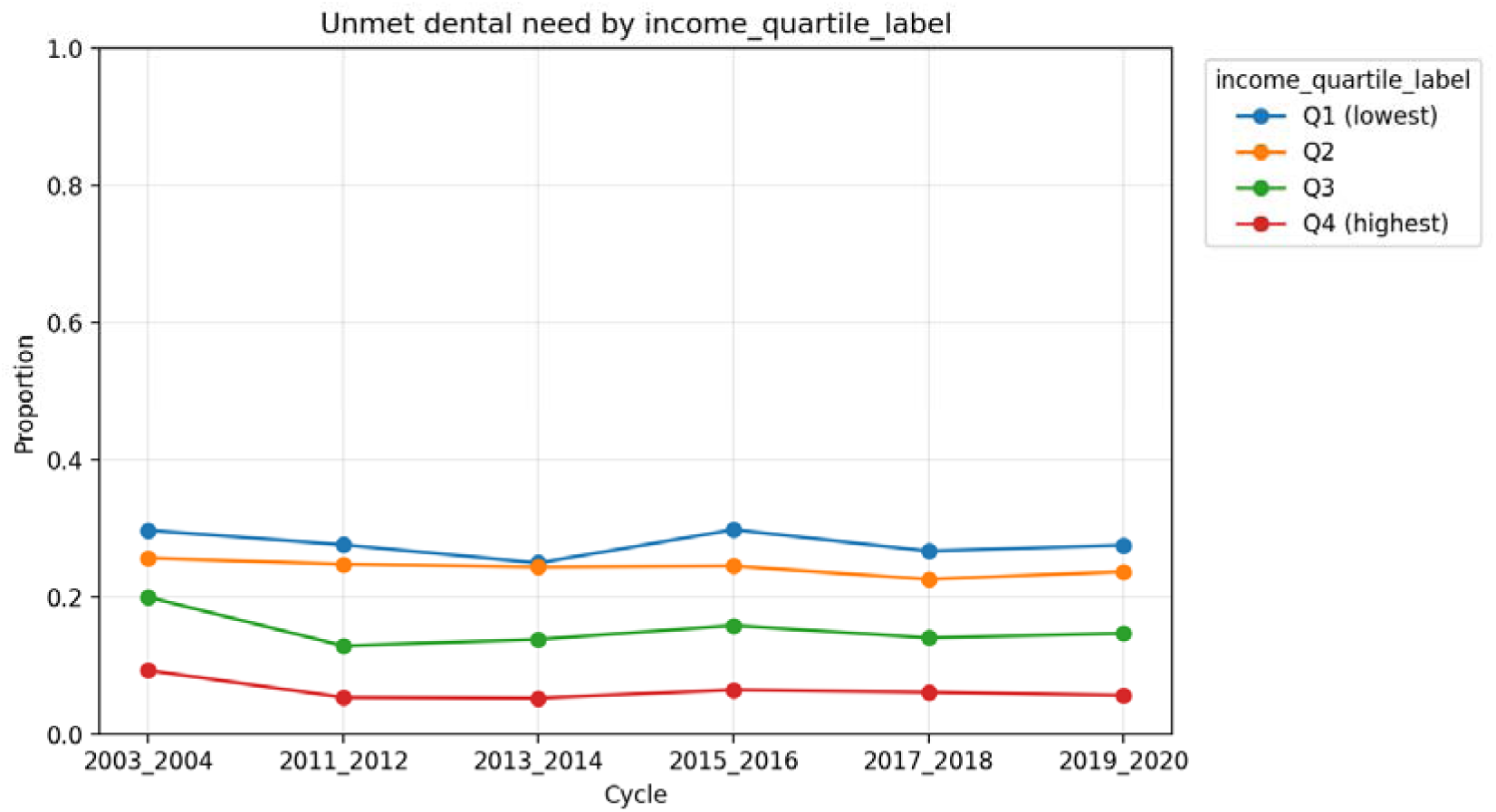

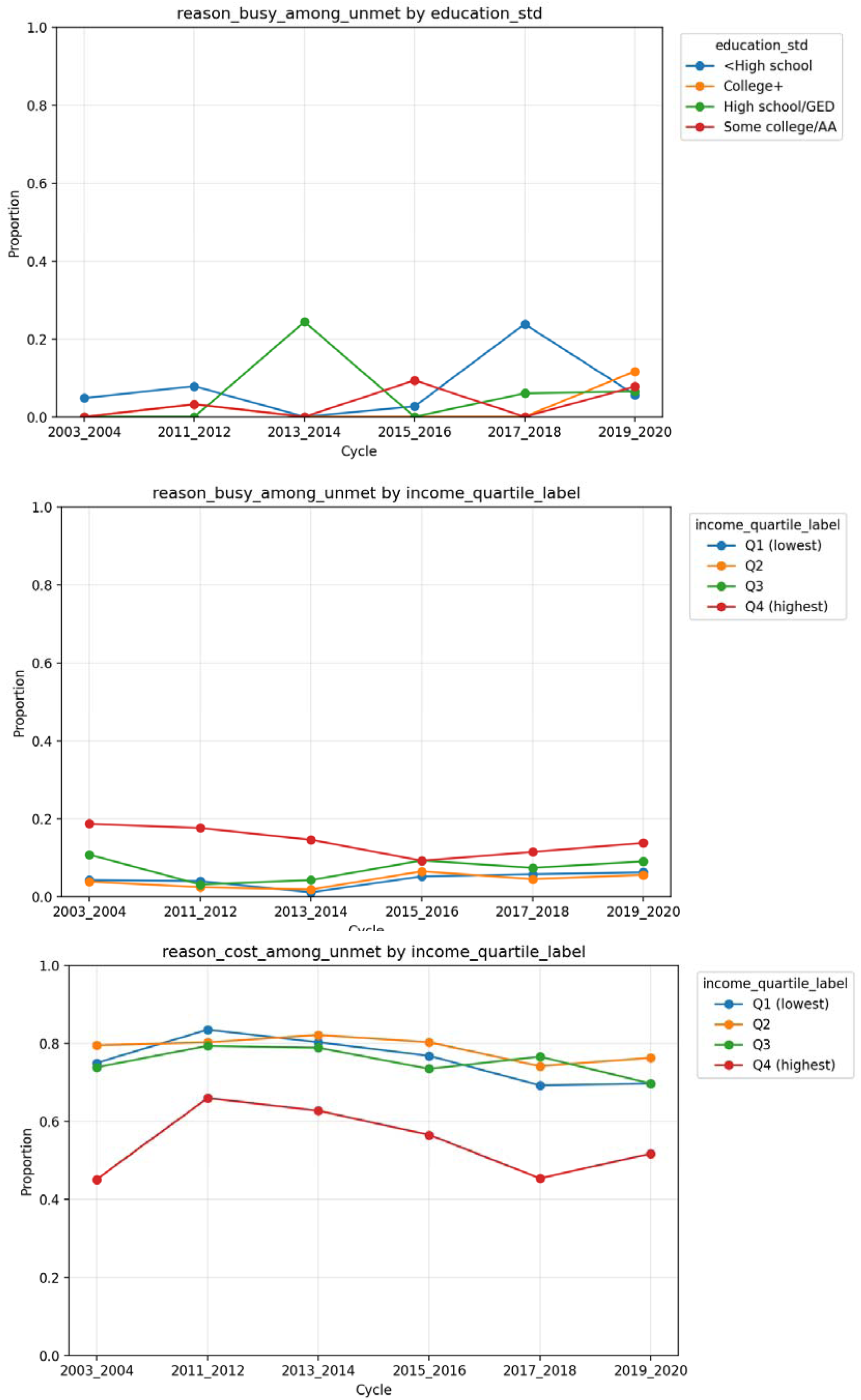

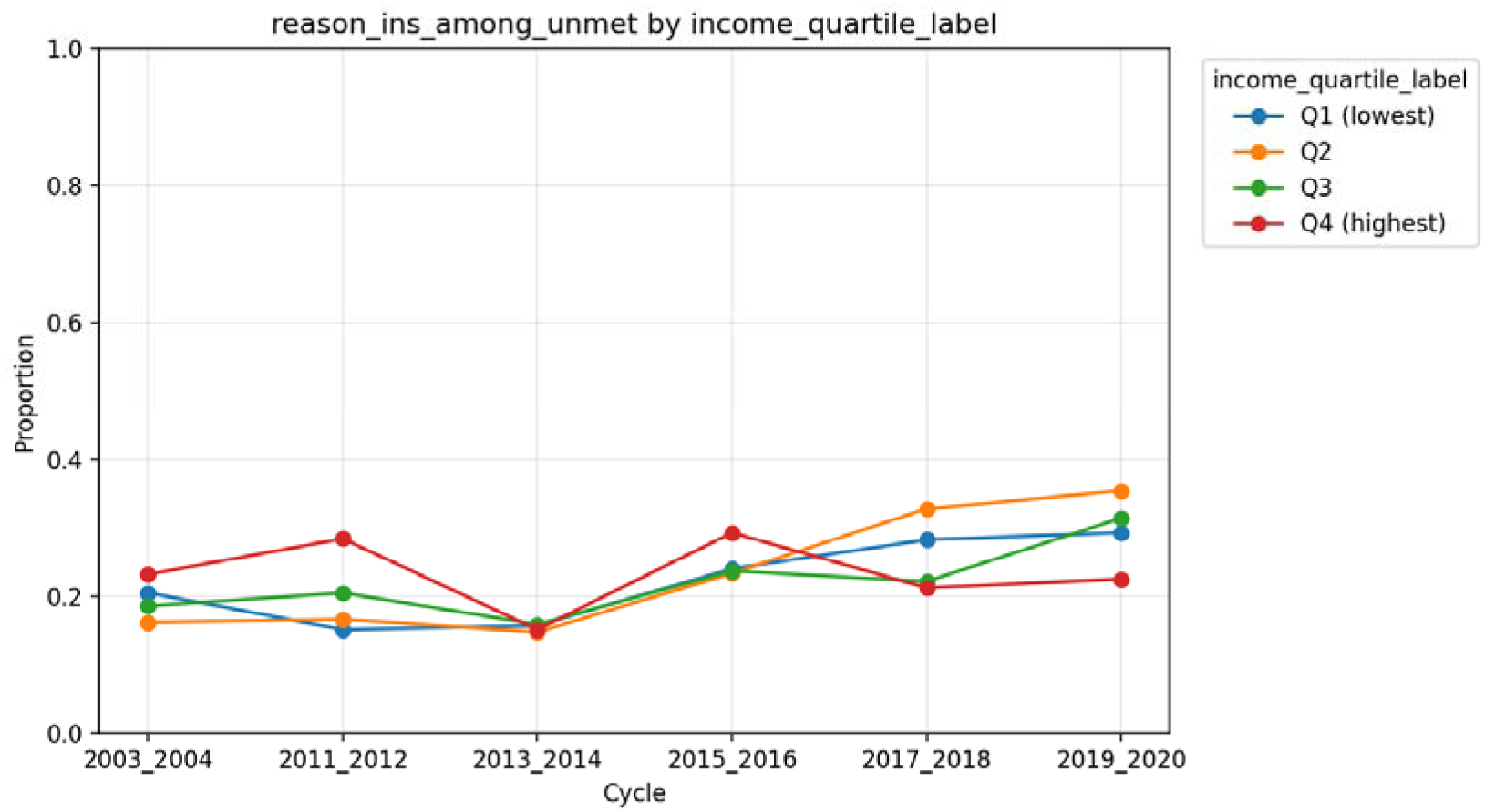

##### 3.1.1.4 Data quality note

It should be noted that the prevalence trend of caries (any_caries) showed a significant shift from 2011-2014 to 2015-2018 (76% to 45%), which is most likely due to methodological changes in testing and should not be considered as evidence of genuine decline. Additionally, proxy data for periodontal disease (perio) exhibited substantial missing values or outliers (e.g., 100%) across multiple cycles, rendering them unreliable. Consequently, the outcome analysis in this study was strictly limited to DMFT and self-reported health.

#### 3.1.2 WLS/Logit Model: Independent Effects of SES on Behavior

After controlling for covariates such as age, sex, race, brushing, and beverage intake, the SES variable maintained strong independent effects on both oral health outcomes (see Table X for specific coefficients).

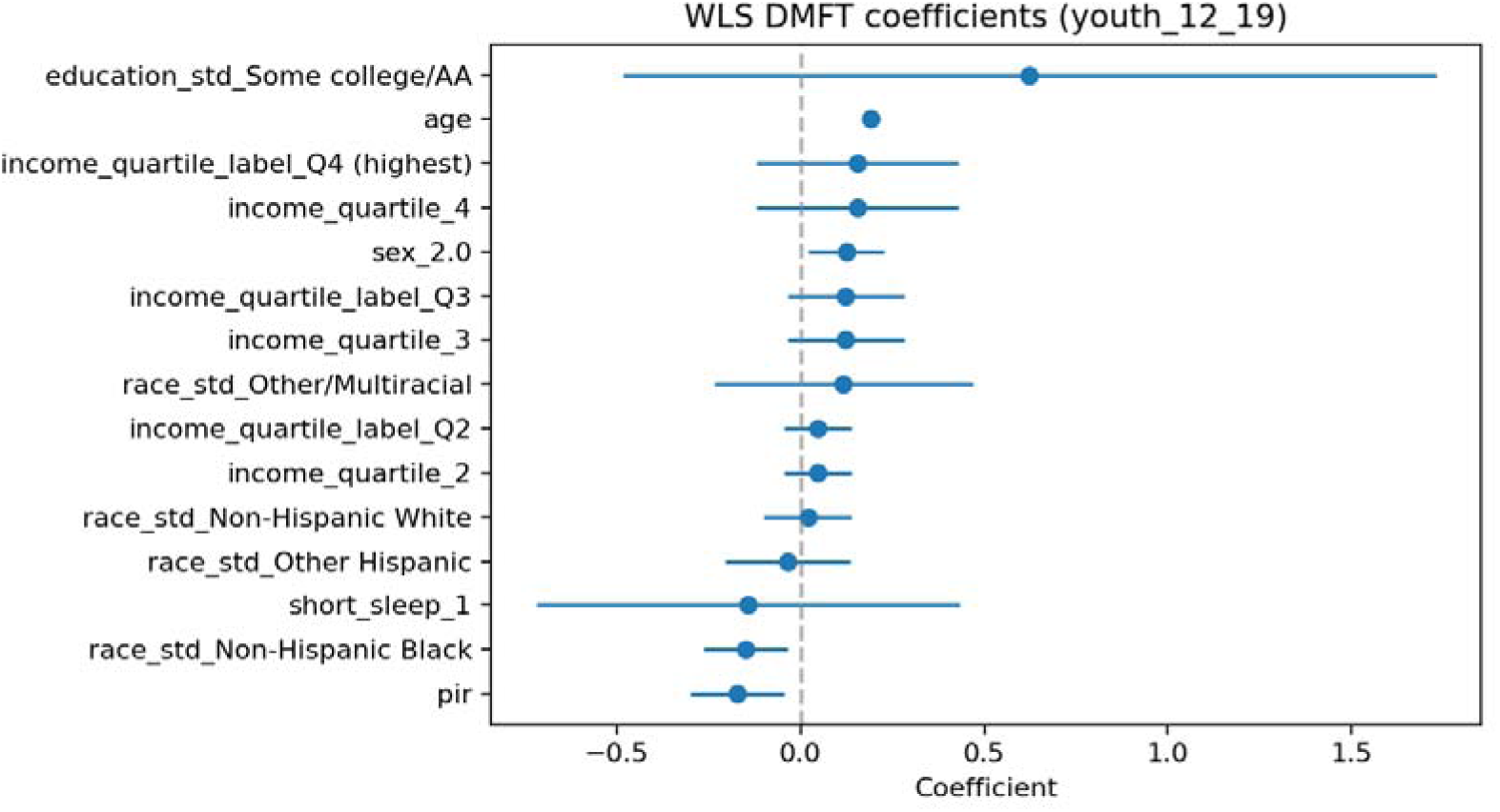

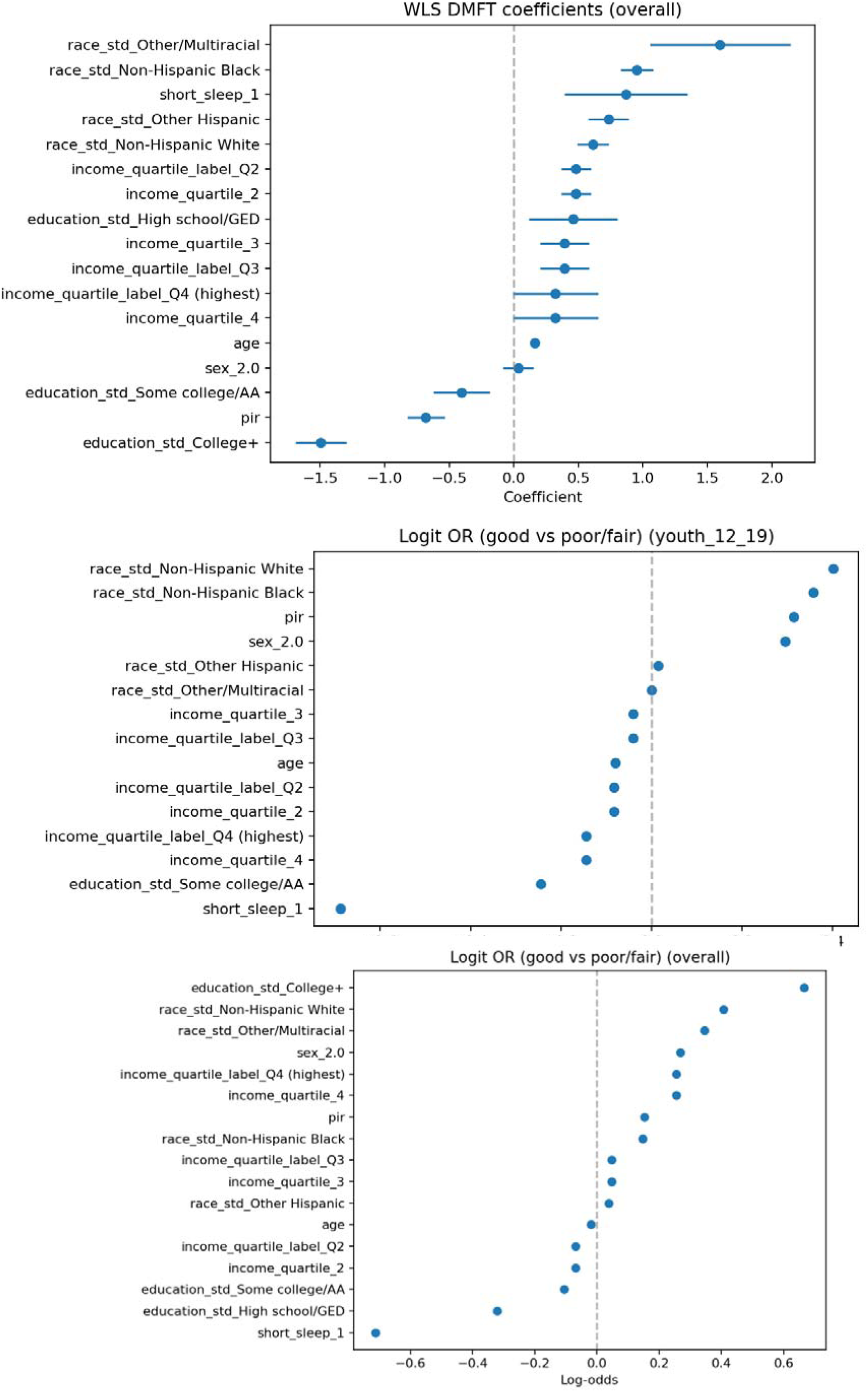

significant negative value (-0.68, p<1e-19), indicating that higher income correlates with lower DMFT. The College+ education coefficient was also significantly negative (-1.49, p<0.05), demonstrating that college education or above serves as a strong protective factor. The age coefficient exhibited a significant positive value (+0.165), consistent with the cumulative exposure pattern of dental caries. Furthermore, both short sleep duration and caffeine intake were positively associated with higher DMFT scores in the WLS (DMFT) model.

##### 3.1.2.2 Logit (self-rated ’good’)

The PIR coefficient was β ≈ ≈β ≈ ≈β ≈ significantly positive (+0.153, p <0.05, OR 1.17), indicating higher self-rated ’good’ probability with higher income. The College+ education coefficient was also significantly positive (+0.667, p <0.05, OR 1.95). The short sleep coefficient was significantly negative (-0.712), suggesting lower self-rated ’good’ probability among short sleepers.

##### 3.1.2.3 Behavioral Factors (PCA)

The independent behavioral model (Behavior_Logit) demonstrated that brushing frequency (OR <1) was a protective factor against caries, while the frequency of consuming sugary beverages (OR> 1) was a risk factor. The principal component analysis (Behavior_PCs_Logit) revealed that PC1 (comprehensive health behaviors) was significantly associated with lower caries risk (OR <1). This provides a biological rationale for subsequently considering behavior as a mediating pathway.

#### 3.1.3 Intermediary Analysis and Robustness

Mediation analysis results (see Table X, mediation_summary.csv) confirmed the mediating role of health care accessibility and sleep behavior between SES and oral health (CI was significant if it did not cross 0).

##### 3.1.3.1 Pathway 1: Medical Access

(1) PIR (income) → accessibility → outcome: The indirect effect of "most recent visit" on DMFT (negative IE) and self-rated "good" (positive IE) was significant. The indirect effect of "unmet need" on self-rated "good" (positive IE) was significant, but not on DMFT.

(2) Education → accessibility → outcome: The indirect effect of "most recent visit" on both DMFT (negative IE) and self-rated "good" (positive IE) was significant. The indirect effect of "unmet need" on self-rated "good" (positive IE) was significant, but not on DMFT.

(3) Interpretation: Higher SES (income and education) significantly improves self-rated oral health by increasing dental visit rates and reducing (subjectively perceived) unmet needs. The rise in visit rates also has a significant indirect effect on reducing DMFT.

##### 3.1.3.2 Pathway 2: Health Behaviors (Sleep)

(1) PIR (income) → sleep → outcome: Both the indirect effects of ’short sleep’ on DMFT (negative IE) and self-rated ’good’ (positive IE) were significant.

(2) Education → sleep → outcome: no significant indirect effect of "short sleep" (IE 0)≈

(3) Interpretation: The effect of income on oral health is partly realized through sleep (high income → less short sleep → better oral health). The link between education and sleep is weaker, indicating that sleep problems may be more related to socioeconomic stress or living environment related to income.

##### 3.1.3.3 Robustness and RCS

Robustness analysis (robustness_summary.csv) shows that the direction and significance of the effects of PIR and high education level (College+) remain stable across different covariate sets and time windows. Restricted cubic spline (RCS) analysis (rcs_*.png) suggests that there is a nonlinear relationship between PIR and age, with the protective effect of PIR being most steep in the low-income range (near the poverty line).

### 3.2 WHO Regional Macro Analysis Results

#### 3.2.1. area differentiation

WHO macro data demonstrate strong geographic clustering in oral health outcomes. Most outcomes (e.g., untreated caries rate, prevalence of severe periodontal disease, and prevalence of anodontia) show statistically significant differences across regions (ANOVA/Kruskal-Wallis, P <0.05). In batch regression models, regional dummy variables serve as strong predictors with high explanatory power.

#### 3.2.2. Economic and Resource Indicators:Correlation and Paradox

The correlation between national economic and resource (per capita GDP, dental density, health expenditure, etc.) and oral health outcomes is complex and even paradoxical in cross-sectional studies.

3.2.2.1 The rate of toothlessness (20+) was strongly positively correlated with *ρ* ≈ ≈ *ρ* ≈≈ the per capita dental expenditure (US$) (+0.668, P 3.3e-26) and the total dental expenditure (million US$) (+0.609, P 6.7e-21). We believe this is not "spending that leads to toothlessness," but rather a classic "structural confounding" in cross-sectional data. Countries with higher spending tend to be those with higher aging, better access to healthcare systems (especially extraction treatments), and more comprehensive reporting systems (often concentrated in specific high-income regions).
3.2.2.2 Severe periodontitis (15+ years) showed no significant correlation with either ρ ≈ ≈ ρ ≈ ≈per capita dental expenditure (-0.021, P=0.772) or total dental expenditure (-0.041, P=0.573). Interpretation: No stable relationship was found in the cross-section, which may be diluted by regional differences, behavioral factors (smoking and alcohol) and heterogeneity of measurement caliber.

##### 3.2.2.3 Untreated caries

There was no significant correlation with either per capita dental expenditure or total dental expenditure, for either deciduous ρteeth (1-9 years) or permanent teeth (5+ years) (values between-0.07 and +0.10, P> 0.15).

Interpretation: Although it is intuitive that better resources should lead to fewer untreated caries (weak negative correlation direction), the evidence is extremely weak on a macro cross-sectional level, reflecting high heterogeneity in regions, policies, behaviours and socioeconomic status.

##### 3.2.2.4 Missing indicator notes

In the enriched data (enriched_data.csv) for this Spearman correlation analysis, the WDI’s GDP/population field and dentist density column failed to merge successfully. Based on empirical judgment and the dominance of regional dummy variables in the OLS model, it is inferred that per capita GDP and dentist density are typically negatively correlated with "untreated caries/periodontal disease," but this effect is often diluted across cross-sections by (age-related, behavioral, and sugar consumption-related) regional structures.

#### 3.2.3. Policy Coverage and Structural Impacts

3.2.2.1 The nominal policy coverage (e.g. whether dental care is included in basic health insurance) is generally not directly correlated with disease prevalence, suggesting that more granular policy intensity and time series data are needed to identify the true effect of policies.
3.2.2.2 Structural factors (e.g. "tropical" or not) are mostly negative for resource input indicators (i.e. tighter resources), and the direction of the relationship between resource input and disease outcomes is influenced by multiple factors, and is not always consistent among different outcomes.

### 3.3 Age-stratified SES gradients and predicted probabilities

3.3.1 Age-stratified models showed that the income gradient in oral health persisted across the life course but varied in magnitude. In linear models for DMFT, the PIR coefficients were negative in all age groups, indicating that higher income was consistently associated with fewer decayed, missing or filled teeth. The estimated coefficients were approximately −0.25 for adults aged 20–44 years, −0.79 for those aged 45–64 years, and −1.07 for adults aged ≥65 years, suggesting that a one-unit increase in PIR is associated with progressively larger absolute reductions in DMFT at older ages. Education-related disadvantage was also evident: low education categories were associated with significantly higher DMFT in all age strata.
3.3.2 In logistic models for self-rated oral health, higher PIR was associated with a higher likelihood of reporting good oral health across all age groups. The odds ratios for PIR were approximately 1.48 (20–44 years), 1.46 (45–64 years) and 1.33 (≥65 years), respectively, while higher education levels independently increased the probability of self-rated good oral health.
3.3.3 Predicted probability curves based on these models (Figure X; predicted_curves.csv, pir_quartile_age_curves.png) further illustrate the SES gradient. Within each age group, the predicted probability of self-rated good oral health increased monotonically from PIR Q1 to Q4. Among adults aged 20–44 years, the predicted probability rose from about 0.60 in Q1 to 0.88 in Q4, whereas among those aged ≥65 years it increased from approximately 0.67 to 0.86 across the same income quartiles. These findings underscore a consistent and substantial income gradient in perceived oral health across the adult life course, with somewhat larger gains associated with higher income in younger and middle-aged adults.

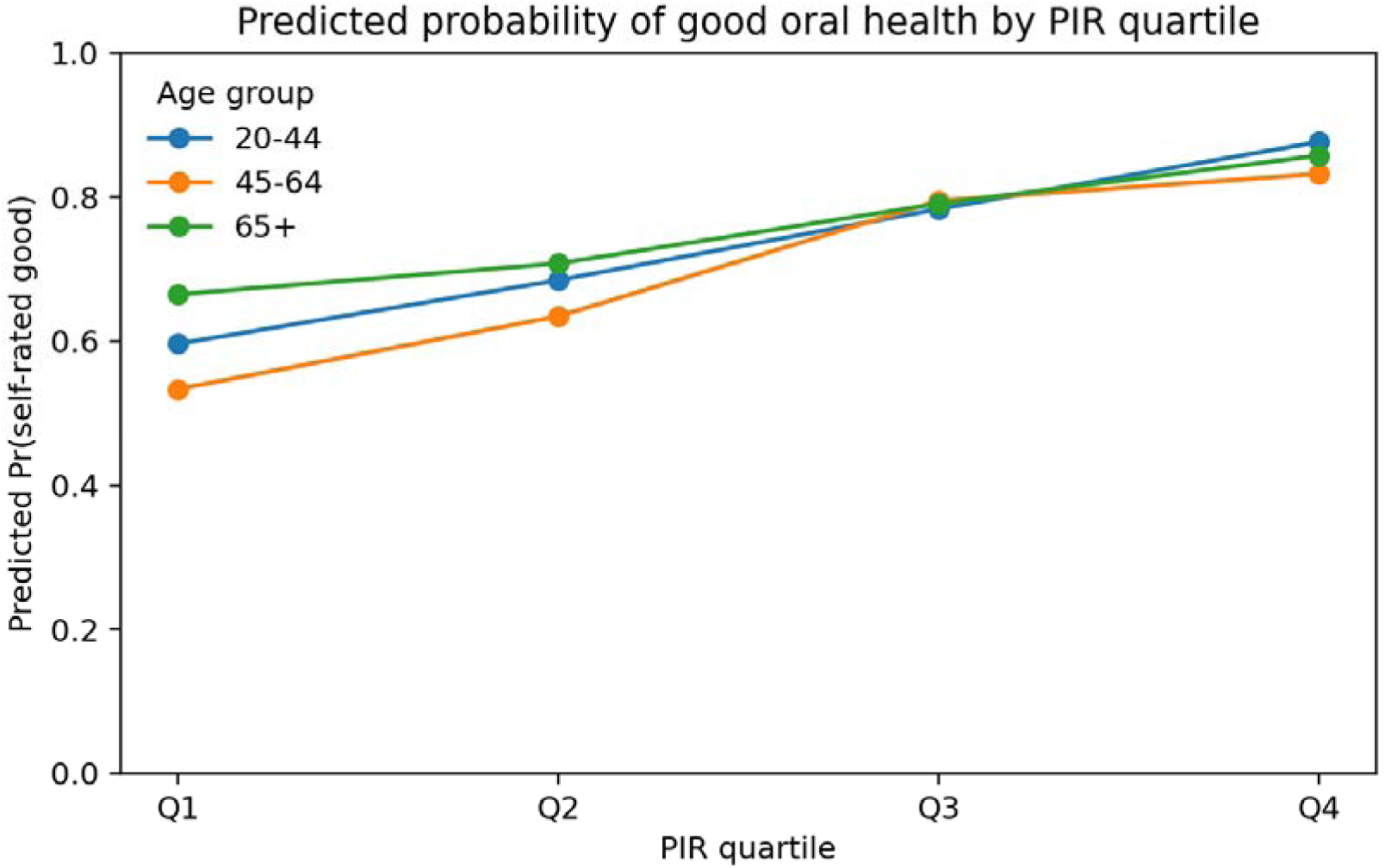

### 3.4 Income-related inequality indices (CI, SII and RII)

3.4.1 Summary inequality indices confirmed that oral health in the United States is strongly patterned by income. The overall concentration index for DMFT was approximately −0.105, indicating that a disproportionate share of caries and tooth loss burden is concentrated among lower-income adults. In contrast, the CI for self-rated good oral health was around +0.094, suggesting that good oral health is concentrated among higher-income groups. Across age strata, the 45–64-year group exhibited the most pronounced inequalities on both measures, consistent with the descriptive and regression findings.
3.4.2 The Slope Index of Inequality (SII) for DMFT indicated that moving from the lowest to the highest income rank was associated with a reduction of about 2.48 teeth affected by decay, missing or fillings on average. For self-rated good oral health, the Relative Index of Inequality (RII) on the odds ratio scale was approximately 8.05, implying that adults at the top of the income hierarchy had about eight times higher odds of reporting good oral health compared with those at the bottom, after accounting for age, sex and race/ethnicity. Together, these indices highlight the magnitude of income-related inequalities in both objective and subjective oral health outcomes, particularly among middle-aged adults.

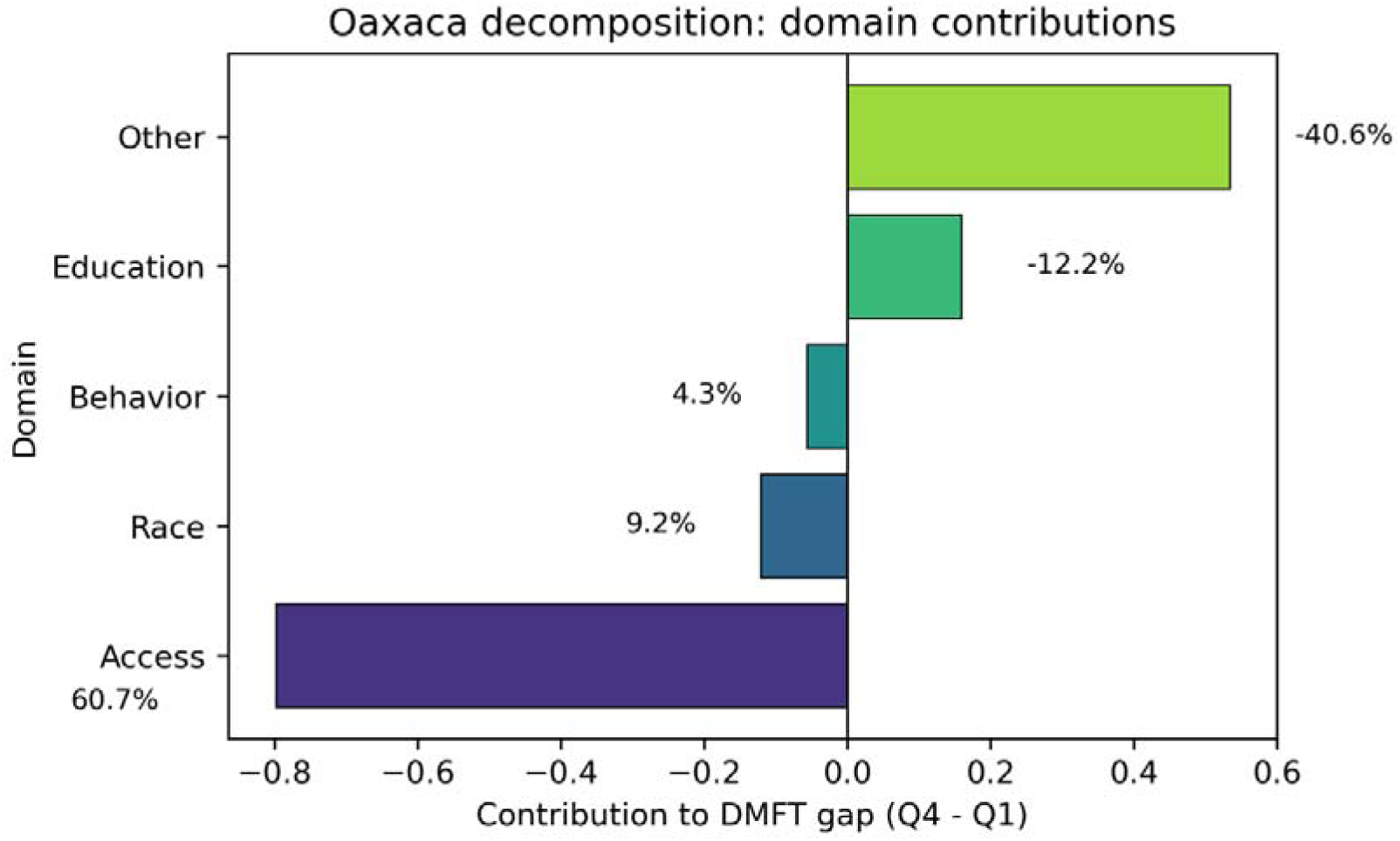

### 3.5 Oaxaca**–**Blinder decomposition of income-related gaps in DMFT

3.5.1 The Oaxaca–Blinder decomposition provided further insight into the proximal determinants of SES inequalities in caries experience. The mean difference in DMFT between adults in the highest (Q4) and lowest (Q1) income quartiles was approximately −1.31, indicating that high-income adults had, on average, 1.31 fewer affected teeth than low-income adults. The decomposition attributed roughly 24.7% of this gap to the “explained” component arising from differences in observed characteristics, while the remaining 75.3% was assigned to the “unexplained” component, reflecting differences in coefficients and unobserved factors.
3.5.2 At the domain level, variables related to access to dental care—primarily annual dental visits and unmet dental needs—played a dominant role. The access domain contributed about −0.88 to the DMFT difference, accounting for roughly 67% of the total gap. In contrast, age (per 10-year increase) contributed a positive 0.55, partially offsetting the access effect, and race/ethnicity variables collectively contributed around −0.11. Detailed tables showed that having had a dental visit in the past year and not reporting unmet dental needs were both strongly protective, and that their more favourable distribution among high-income adults explained a substantial portion of the observed inequality in DMFT. These findings support the interpretation that inequalities in access to dental care are a central, measurable pathway through which SES shapes cumulative caries experience, although a large part of the SES gap remains unexplained by observed variables.

### 3.6 State-level Medicaid expansion, adult dental benefits and state-level outcomes

3.6.1 In the state-level difference-in-differences analysis, Medicaid expansion and adult dental benefit generosity were not strongly associated with changes in aggregate oral health outcomes over the study period. In models with state and year fixed effects and standard errors clustered at the state level, the coefficient on the Medicaid expansion indicator for dental visit rates among adults was −0.0399 (standard error [SE] 0.0352; p = 0.26), and the coefficient on the ordinal measure of adult dental benefit generosity was −0.0418 (SE 0.0378; p = 0.27). The model explained about 21.9% of the variance in state-level dental visit rates (R² = 0.219).
3.6.2 For any tooth loss and complete edentulism among adults aged ≥65 years, effect estimates were also modest and statistically non-significant. The Medicaid expansion indicator was associated with a change of −0.012 in the prevalence of any tooth loss (SE 0.034; p = 0.73) and +0.241 for complete edentulism in older adults (SE 0.334; p = 0.47). The corresponding coefficients for adult dental benefit generosity were approximately +0.008 for any tooth loss and −0.125 for edentulism among older adults, neither reaching conventional levels of statistical significance.

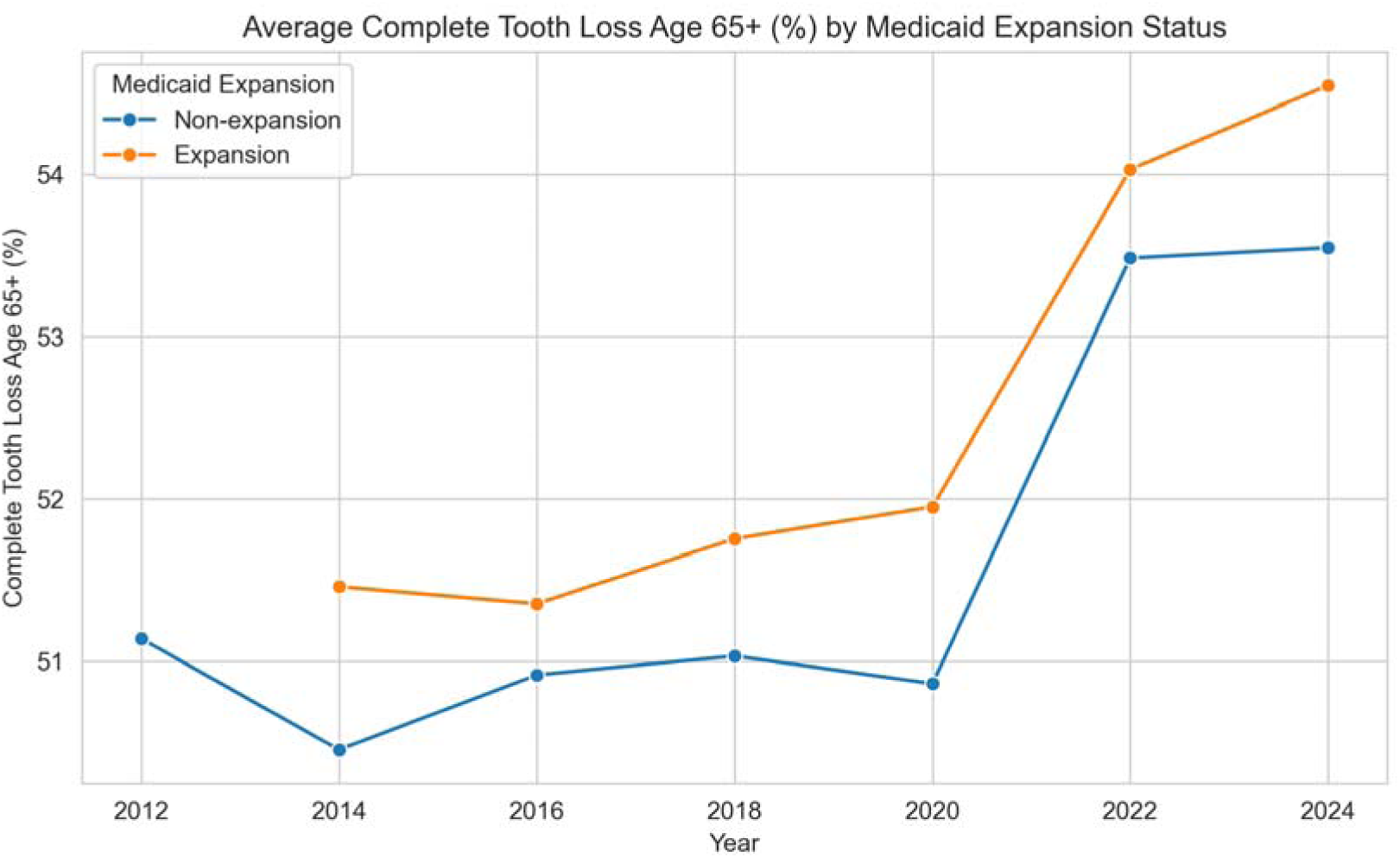

3.6.3 These results suggest that, when examined at the level of overall state populations and aggregate outcomes, recent changes in Medicaid eligibility and adult dental benefits did not produce large, precisely estimated shifts in dental visit rates or tooth loss. This pattern is consistent with the notion that tooth loss and edentulism are highly cumulative, distal outcomes that respond slowly to policy changes, and that policy effects concentrated among low-income Medicaid beneficiaries may be diluted in state-level averages that include large numbers of individuals not directly affected by Medicaid policy.

## 4. Discussion

### 4.1 Domestic and international comparisons, and mechanisms of health inequity

#### 4.1.1 Comparison between Macro (WHO) and Local (NHANES)

This study combines WHO macro data and NHANES micro data to provide multi-level evidence for oral health inequalities. WHO data reveals the "regional patterns" of global inequalities and the "complexity" and "paradox" of macro correlations, while NHANES data opens the "mechanistic black box" of intra-national inequalities.

One of the key findings of this study is the exposure contrast between macro-level correlations (WHO) and local mechanisms (NHANES). At the WHO macro level, economic ρ ≈indicators (dental expenditure) and health outcomes (dental free rate) exhibit a paradoxical +0.67 positive correlation. This clearly highlights the risk of an "ecological fallacy": on a macro level, countries with aging populations, higher dental extraction preferences, and well-established reporting systems (often high-expenditure countries) cluster together, creating this confounding effect. While this macro-level correlation appears "real" as it reflects inter-country variations in population structure and healthcare systems, misapplying it to individual levels may lead to the paradox of "this batch of teeth causing tooth loss."

Conversely, across all NHANES cohorts, we observed a strong negative correlation (-0.68) between SES (PIR) and DMFT after β ≈ ρ ≈controlling for key confounding factors like age. This contrasts with the argument that macro-level correlations (or lack thereof, such as savings versus interruption risk) cannot address local realities. To understand health status, we must delve into individual monitoring levels, as macro data can only provide context and cannot replace local populations.

The "economic barriers" identified in this NHANES study form the core of the leadership sequence and provide a potential explanation for the global significance mechanism. At the global level, national economic/resource indicators such as per capita GDP and dental clinic density are associated with oral health outcomes. The findings suggest that this macro-level correlation is likely mediated by "payment capacity" (PIR) and "service accessibility" (visit rates and unmet demand) across various levels.

Beyond average associations, our age-stratified analyses and inequality indices highlight a distinct life-course pattern of oral health inequalities. The income coefficient for DMFT became more negative with age, and SII/RII estimates suggested that middle-aged adults (45–64 years) bear the steepest socioeconomic gradients in both objective and subjective oral health. The CI, SII and RII results complement our regression findings by showing that not only are poor outcomes concentrated among low-income groups, but the relative advantage enjoyed by high-income adults in self-rated oral health is substantial, with an eight-fold difference in the odds of reporting good oral health between the extremes of the income distribution. These findings reinforce the view that oral health inequalities are not confined to early life or old age, but are most pronounced in midlife, when cumulative exposures to adverse working conditions, stress and limited access to care may converge.

### 4.2 The Interaction of Economy, Behavior and Institution

The analysis of this study clearly demonstrates the roles of economic, behavioral, and institutional factors, drawing on an understanding of the SDOH theoretical framework.

4.2.1 The socio-psychological economic-behavioral pathway (PIR → short sleep → oral health) exemplifies the classic theory of ’biological manifestation of socioeconomic status.’ It demonstrates how economic stress (low PIR) leads to unhealthy lifestyle choices (short sleep), ultimately causing severe health damage ^[17]^.The low PIR group may be more likely to work shift shifts, be in chaotic financial environments, or face greater stress, all of which have direct social impacts on sleep deterioration, potentially damaging oral health through immune regulation or increased reactive responses ^[18]^. Interestingly, the link between education and sleep is weaker, suggesting that income (material resources) may play a more direct role in buffering stress and rest than education (knowledge).
4.2.2 The economic system pathway (material and service): SES → healthcare accessibility → oral health. This pathway confirms the critical role of institutional factors (health insurance, service pricing) in moderating the impact of SES ^[19,20]^.RCS analysis reveals that the effect of PIR is steep at the lower end of the income scale, which has important policy implications. It indicates that near the poverty line, a small increase (or decrease) in income has the greatest marginal effect on oral health. Measures aimed at improving the economic capacity of the most affluent groups (such as raising the minimum wage or expanding tax credits) may have the greatest impact on improving oral health.

#### 4.2.3 Crossing the Gap between Knowledge and Practice

The findings of this study validate the concepts of "intersectionality" and the "knowledge-action gap." Groups are trapped in a vicious cycle: low income (material) triggers high stress (psychological) and low employment rates (institutional), which in turn leads to unhealthy behaviors (e.g., sleep deprivation) and poor health outcomes. As prior research indicates, health school education programs may effectively enhance knowledge dissemination (knowledge), but when students’ families cannot afford treatment costs or when local communities lack dentists (a "resource mismatch" in social terms), such knowledge fails to translate into health improvements. The study identifies "economic barriers" as the primary obstacle, reaffirming that bridging the knowledge-action gap hinges not on information transmission, but on systemic institutional and economic constraints.

#### 4.2.4 The enlightenment of the two-tailed model

The significance lies in the fact that ’unmet needs’ exert a stronger influence on ’self-rated health’ than DMFT. This suggests that ’unmet needs’ themselves represent a perception of social injustice and systemic inequity in healthcare, which directly undermines individual health. In contrast, DMFT serves as a monitored, lifelong clinical indicator whose changes are slower and more dependent on ’actual treatment’ rather than ’perceived unmet needs.

### 4.3 Policy Implications and the Path to Oral Health Equity

Based on the above findings, this study provides important implications for public health policy, which lies in the shift from "individual responsibility" to "social and institutional responsibility".

#### 4.3.1 Going beyond "health education"

Health education for low-income groups (such as "brushing teeth regularly") is not sufficiently attention-grabbing and may even reinforce the tendency toward "oral health problems." NHANES data shows that the reason they do not seek medical care is "unaffordable".

#### 4.3.2 Addressing economic barriers (core)

To improve oral health levels and address the economic accessibility issues of essential dental care. This includes expanding mandatory coverage of dental care for Medicaid adults in the United States; and exploring the inclusion of key dental prevention services (such as fluoride application, fissure sealants) and dental procedures (such as fillings) in the basic medical insurance reimbursement list in China.

#### 4.3.3 Upstream prevention and community projects

School oral health programmes should go beyond intermittent forums and evolve into long-term, comprehensive hazard models that provide preventive services (e.g. regular fissure sealants and full mouth fluoride) and facilitate enhanced interaction with parents.At the same time, public policies aimed at reducing universal sugar consumption (e.g. a tax on sugar-sweetened beverages) and ensuring that all people have access to effective fluoride toothpaste ^[8]^.

#### 4.3.4 Focus on social determinants (upstream)

The finding of sleep as an important factor reminds us that oral health is part of overall health and the social environment. Improving working conditions for low-income people (e.g. reducing the inability to work shifts), reducing their life stress (e.g. providing hypertension), and improving their living environment (e.g. reducing pollution and noise) are important to improve their overall health, including oral health.

Our state-level difference-in-differences analyses did not detect large, statistically significant average effects of the ACA Medicaid expansion or measured variation in adult Medicaid dental benefit generosity on aggregate dental visit rates or tooth loss outcomes. These null findings should be interpreted with caution. First, the outcomes we examined at the state level—overall dental visit rates and tooth loss/edentulism—are distal and highly cumulative, and are likely to respond slowly to policy changes that occurred mainly during the 2010s. Second, the state-level prevalence estimates reflect all adults, including many who are not Medicaid-eligible; any benefits concentrated among low-income enrollees may therefore be diluted when averaged across whole state populations. Third, our policy measures, although based on established classifications, may not fully capture within-state variation in benefit design, provider participation, reimbursement levels and implementation.

Taken together, these considerations suggest that the DID results should be seen as evidence against large, short-term average effects on state-wide aggregate outcomes, rather than as proof that Medicaid dental policies are irrelevant for oral health. In fact, prior studies using individual-level data have reported clear impacts of Medicaid expansion and adult dental benefit enhancements on dental visit rates, unmet dental needs, and emergency department use for dental conditions among low-income adults. In this context, our micro-level NHANES analyses, which demonstrate a strong SES gradient and a central role for access to care, are more sensitive for identifying mechanisms of inequality than the state-level analyses focusing on distal endpoints.

The contrast between our strong and consistent SES gradients in NHANES and the modest, imprecise policy effects observed in state-level DID models carries an important policy message. Expanding public coverage for dental care—through Medicaid or similar schemes—is likely a necessary but not sufficient condition for reducing oral health inequalities. Coverage expansion can alleviate financial barriers and reduce cost-related unmet need, as suggested by prior literature, but our results indicate that much of the inequality in DMFT and self-rated oral health remains linked to broader income and education inequalities and to upstream social determinants such as working conditions, stress and sleep. Policies that seek to improve oral health equity must therefore combine improvements in dental coverage and service delivery with broader social policies that address poverty, educational disadvantage and the social patterning of health-related behaviours. This conclusion is relevant not only for the United States but also for China and other countries seeking to integrate oral health into universal health coverage agendas.

## 5. Conclusion and Recommendations

This study, through a multilevel analysis of WHO macro data and NHANES micro data, confirms significant socioeconomic disparities in oral health both globally and within the United States.

The correlation between macroeconomic indicators and health outcomes is often obscured by structural factors like ρ ≈aging, creating a paradox (e.g., higher expenditures correlate with higher toothlessness rates, +0.67). However, micro-level mechanisms within countries clearly demonstrate that socioeconomic status influences individuals’ economic accessibility to dental services (economic barriers) and health-related behaviors such as sleep patterns, which ultimately translate into oral health outcomes.

Future policy development should transition from "health education" to "barrier removal", prioritizing solutions for low-income populations’ healthcare affordability, actively addressing social determinants of health, and improving geographical equity in medical resource distribution. Specifically, policies should tackle both "price barriers" (e.g., insurance coverage) and "time barriers" (e.g., expanding community service points and streamlining procedures). By integrating oral screening into primary care, we can bridge the gaps in preventive care caused by insufficient public education and the pain-driven approach, thereby promoting substantive equity in oral health.

## 6. Limitations

The study has the following limitations:

1. Approximate Estimation (NHANES): The Public Health Standard Error of the Unit (PSU-robust) used in this study is an estimate derived from NHANES’ complex error design.
2. Cross-sectional design (NHANES, WHO): All data are cross-sectional, making it impossible to establish strict causality. While the model theoretically follows a temporal sequence (SES → inclusion → causality), reverse causality may occur. For instance, severe oral issues (e.g., multiple tooth defects) could lead to speech impairment and aesthetic concerns, thereby reducing an individual’s bargaining power in the labor market and resulting in lower income (PIR).
3. Data quality limitations (NHANES): The NHANES data proxy for caries (any_caries) and periodontal disease (perio) was unreliable in some cycles, thus the study’s conclusions regarding DMFT and self-rated health were strictly limited.
4. Method bias: The measurement tools of large-scale epidemiological surveys (e.g. NHANES) differ from clinical accurate diagnosis. For example, some studies have indicated that the CPI method based on exponential teeth may systematically underestimate the true prevalence of periodontal disease, and the DMFT measurement in this study may also have similar survey bias.
5. Missing variables (NHANES): The urban-rural disparity we initially aimed to investigate could not be addressed due to public data gaps (URDHHLOC), representing a critical dimension of omission.
6. Missing factors (environment): The study focused on SES structure and did not identify other fundamental physical environmental factors, such as endemic fluorosis, which represents another SES-dependent longitudinal pathway.
7. Covariance of variables (NHANES): PIR and income quartile labels are highly collinear. Therefore, PIR is used as the main income explanatory variable in the main model, and the two variables should not be input into the model simultaneously.
8. Ecological fallacy (WHO): Macro-level conclusions (at the national level) cannot be directly extrapolated to individual cases. The WHO analysis paradox of ’a normal distribution between expenditure and toothlessness rate’ is a classic example, reflecting structural differences between countries (e.g., aging populations, treatment models) rather than individual causality.
9. Data quality limitations (WHO): The macro analysis relied on data quality reported from various regions, which showed significant heterogeneity. Furthermore, some key variables (e.g., dental clinic density) were not successfully merged in this enrichment analysis, limiting the scope of correlation analysis.

## Data Availability

l data produced in the present study are available upon reasonable request to the authors

https://www.cdc.gov/nchs/nhanes/index.html

https://www.who.int/data/collections

https://www.kff.org/medicaid/status-of-state-medicaid-expansion-decisions/

https://www.chcs.org/media/Medicaid-Adult-Dental-Benefits-Overview-Appendix_091519.pdf

https://www.carequest.org/Medicaid-Adult-Dental-Coverage-Checker

https://www.macpac.gov/subtopic/medicaid-expansion/

## Ethics and Reporting

This study relied exclusively on secondary analysis of existing, publicly available data and did not involve any new intervention or contact with human participants or animals. Individual-level microdata were obtained from the National Health and Nutrition Examination Survey (NHANES), and state-level prevalence estimates were drawn from the Behavioral Risk Factor Surveillance System (BRFSS). Both NHANES and BRFSS are conducted by the U.S. Centers for Disease Control and Prevention (CDC); their protocols are reviewed and approved by the National Center for Health Statistics (NCHS) Ethics Review Board (ERB) or equivalent CDC institutional review boards to ensure that participants’ rights and welfare are protected in accordance with U.S. federal regulations. Written informed consent is obtained from all NHANES participants.

In addition, we used aggregated, non-identifiable country- or state-level data from the WHO Global Oral Health and Global Burden of Disease (GBD) platforms, World Bank indicators, and publicly available Medicaid policy sources (e.g. Kaiser Family Foundation, CHCS, MACPAC, CareQuest). These data sources do not contain personal identifiers. Because all datasets analysed in this study are de-identified and publicly accessible, and no new experiments involving human participants were conducted, the present analysis was considered exempt from additional institutional ethics review. The study complies with the principles of the Declaration of Helsinki where applicable and follows STROBE recommendations for reporting observational studies.

## Notes

### Competing Interest Statement

The authors have declared no competing interest.

### Funding Statement

This study did not receive any funding

### Author Declarations

This study relied exclusively on secondary analysis of existing, publicly available data and did not involve any new intervention or contact with human participants or animals. Individual-level microdata were obtained from the National Health and Nutrition Examination Survey (NHANES), and state-level prevalence estimates were drawn from the Behavioral Risk Factor Surveillance System (BRFSS). Both NHANES and BRFSS are conducted by the U.S. Centers for Disease Control and Prevention (CDC); their protocols are reviewed and approved by the National Center for Health Statistics (NCHS) Ethics Review Board (ERB) or equivalent CDC institutional review boards to ensure that participants rights and welfare are protected in accordance with U.S. federal regulations. Written informed consent is obtained from all NHANES participants.In addition, we used aggregated, non-identifiable country- or state-level data from the WHO Global Oral Health and Global Burden of Disease (GBD) platforms, World Bank indicators, and publicly available Medicaid policy sources (e.g. Kaiser Family Foundation, CHCS, MACPAC, CareQuest). These data sources do not contain personal identifiers. Because all datasets analysed in this study are de-identified and publicly accessible, and no new experiments involving human participants were conducted, the present analysis was considered exempt from additional institutional ethics review. The study complies with the principles of the Declaration of Helsinki where applicable and follows STROBE recommendations for reporting observational studies.

### Summary of Updates

New analysis added,include BRFSS policy survey on America

